# No cognitive or psychological impact from returning research Alzheimer disease biomarkers: A delayed-start, noninferiority, randomized clinical trial

**DOI:** 10.64898/2026.05.22.26353881

**Authors:** Sarah M. Hartz, Sacha Jackson, Tammie L.S. Benzinger, Laura Bierut, Alissa Evans, Spondita Goswami, Brian A. Gordon, Jason Hassenstaab, Lisa Hayibor, Erin Linnenbringer, John C. Morris, Krista L. Moulder, Amy Oliver, Lingwei Sun, Suzanne E. Schindler, Chengjie Xiong, Jessica Mozersky

## Abstract

**Importance:** Little is known about the impact of returning Alzheimer disease (AD) biomarkers to cognitively unimpaired (CU) research participants.

**Objective:** Determine whether return of research results (RoRR) negatively impacts longitudinal symptoms of depression and cognition.

**Design:** Randomized, noninferiority, delayed-start clinical trial, 2021-2025

**Setting:** AD biomarker research results offered to CU participants in a longitudinal cohort of community-dwelling older adults.

**Participants:** 341 CU research participants age ≥65 with available biomarkers (*APOE* genotype and either plasma Aβ42/40 or amyloid PET and MRI hippocampal volume) were recruited.

**Intervention(s) (for clinical trials) or Exposure(s) (for observational studies):** Participants were offered their research AD biomarker results (RoRR) with an estimated 5-year risk of symptomatic AD. After consenting, participants were randomized to either receiving results within several weeks (RoRR arm) or 1 year later (delayed-start arm).

**Main Outcome(s) and Measure(s):** Longitudinal change in Geriatric Depression Scale (GDS), Clinical Dementia Rating® sum of boxes (CDR-SB), and global cognitive composite. Outcomes were measured at annual assessments for a longitudinal study of aging.

**Results:** 147 participants received results after randomization: 70 in RoRR arm (average age 75, 60% female), 66 in delayed-start arm (average age 73, 53% female). The observed changes in annual measures did not differ between arms in both those with elevated amyloid (Aβ+) and in those without elevated amyloid (Aβ-) for GDS (Aβ+ difference 0.7, 95% CI 0.0-1.3; Aβ-difference −0.1, 95% CI −0.7-0.5; clinically significant decline >4.0), CDR-SB (Aβ+ difference 0.0, 95% CI −0.1-0.1; Aβ-difference 0.0, 95% CI 0.0-0.1; clinically significant decline >0.5), and cognitive composite (Aβ+ difference −0.10, 95% CI −0.25-0.06; Aβ-difference - 0.05, 95% CI −0.17-0.07; clinically significant decline < −0.26). Secondary analyses found no evidence of association between RoRR and proximity to follow-up testing.

**Conclusions and Relevance:** In the first randomized, delayed-start clinical trial of returning AD research results to CU older-adult participants, no effect was seen on longitudinal changes in symptoms of depression or cognition. This supports evidence that there are no harms to returning AD research results, although the results may not apply to more diverse populations not included in this study.

**Trial Registration:** NCT04699786

**Key points:** *Question:* Does return of Alzheimer’s disease (AD) research biomarker results to cognitively unimpaired older adults negatively impact longitudinal symptoms of depression and cognition?

*Findings:* In this randomized, delayed-start clinical trial, no effect of returning research AD biomarkers was seen on longitudinal changes in symptoms of depression or cognition.

*Meaning:* This supports evidence that there are no harms to returning AD research results, although the results may not apply to more diverse populations not included in this study.

## Introduction

The availability of disease-modifying therapies (DMTs) for Alzheimer disease (AD) and advocacy for return of research results (RoRR) are creating heightened desire for RoRR in AD research studies.^1,2^ However, lack of data regarding the implications of returning research results in the absence of clinical actionability continues to be a significant barrier to returning these results.^2^

A primary concern around disclosing AD research results to cognitively unimpaired older adults is the possibility of negative psychological impact such as increased anxiety or depression.^3,4^ While some studies have found low levels of psychological impact of receiving AD biomarker results,^5^ others, including a meta-analysis, found no short-term psychological impact of disclosing AD biomarker results.^6–9^ In addition, longitudinal studies have found that negative psychological impact of receiving results wanes over time.^6,10,11^

An additional concern is that returning results would have an effect on participants’ cognitive testing results, potentially impacting primary outcomes of the parent research study.^12^ Specifically, learning of an increased risk for AD could cause anxiety that interferes with performance on memory and thinking tests. Only two studies have evaluated this question to date, and both evaluated the impact of learning genetic results on cross-sectional cognitive test results. The first study found that older adults who learned that they carried an *APOE* ε4 allele judged their memory more harshly and performed worse on an objective verbal memory test than did *APOE* ε4+ adults who did not know their results.^13^ A second study compared cognitive performance between three groups of participants who carried autosomal dominant AD mutations: those who knew their mutation status prior to the study onset, those who learned their mutation status during the approximately 3-year study, and those who did not know their mutation status.^14^ Although those who knew their mutation status before the study did not perform worse than those who never learned their mutation status, participants who learned they were mutation carriers during the study had lower scores on a cognitive composite relative to mutation carriers that did not know their status.^14^ This suggests that the negative cognitive impact of learning genetic results on cognitive performance may diminish over time. It is important to note that neither study randomized participants to learn results or evaluated change in cognition over time. In addition, no study has evaluated the impact of amyloid disclosure on longitudinal cognitive outcomes.

To address this gap, we conducted a randomized, delayed-start noninferiority clinical trial to specifically test the impact of learning AD research biomarker results (*APOE* ε4 and amyloid PET or plasma amyloid results) among older adults participating in a longitudinal study of aging that receive annual evaluations including measures of depressive symptoms and cognitive testing.^15^

## Methods

### Trial Design and Oversight

This was a randomized clinical trial conducted at the Charles F. and Joanne Knight Alzheimer Disease Research Center (ADRC) at Washington University School of Medicine in St. Louis, Missouri from January 2021 to October 2025. This trial was conducted in accordance with the principles of the Declaration of Helsinki and the International Conference on Harmonization Good Clinical Practice guidelines. This trial followed the Consolidated Standards of Reporting Trials (CONSORT) reporting guideline. The trial protocol was approved by the Washington University Institutional Review Board, was previously published,^15^ and was registered with clinicaltrials.gov (NCT04699786). All participants gave electronic or written informed consent. An external safety officer was sent semi-annual reports and notified of any adverse events. The study was designed with input from Knight ADRC participants. Participants were allowed to withdraw from the study at any time.

### Participants

Memory and Aging Project (MAP) is a longitudinal study of aging that includes annual clinical and cognitive assessments and periodic biomarker measurements including *APOE* genotyping, brain MRI imaging, amyloid PET scans, and plasma testing of amyloid-related biomarkers. Eligible MAP participants were cognitively unimpaired (defined by Clinical Dementia Rating® (CDR®)^16^ score of 0, as assessed in the past 12 months), aged 65 or older, consented for contact for additional studies, and had research biomarkers available for return.

After processing the biomarkers, eligible participants were mailed a packet of information including a cover letter describing the study with contact information, an educational brochure regarding AD biomarker results and the decision to receive results, and an example consent form (**Figure 1**).^15,17^ If a participant did not contact the study team within 2 weeks, the participant received two follow-up calls to make sure that they received the information and to ask whether they were interested in participating. If interested, an appointment was made with a study coordinator to discuss the study in more detail and give informed consent.

**Figure 1:**
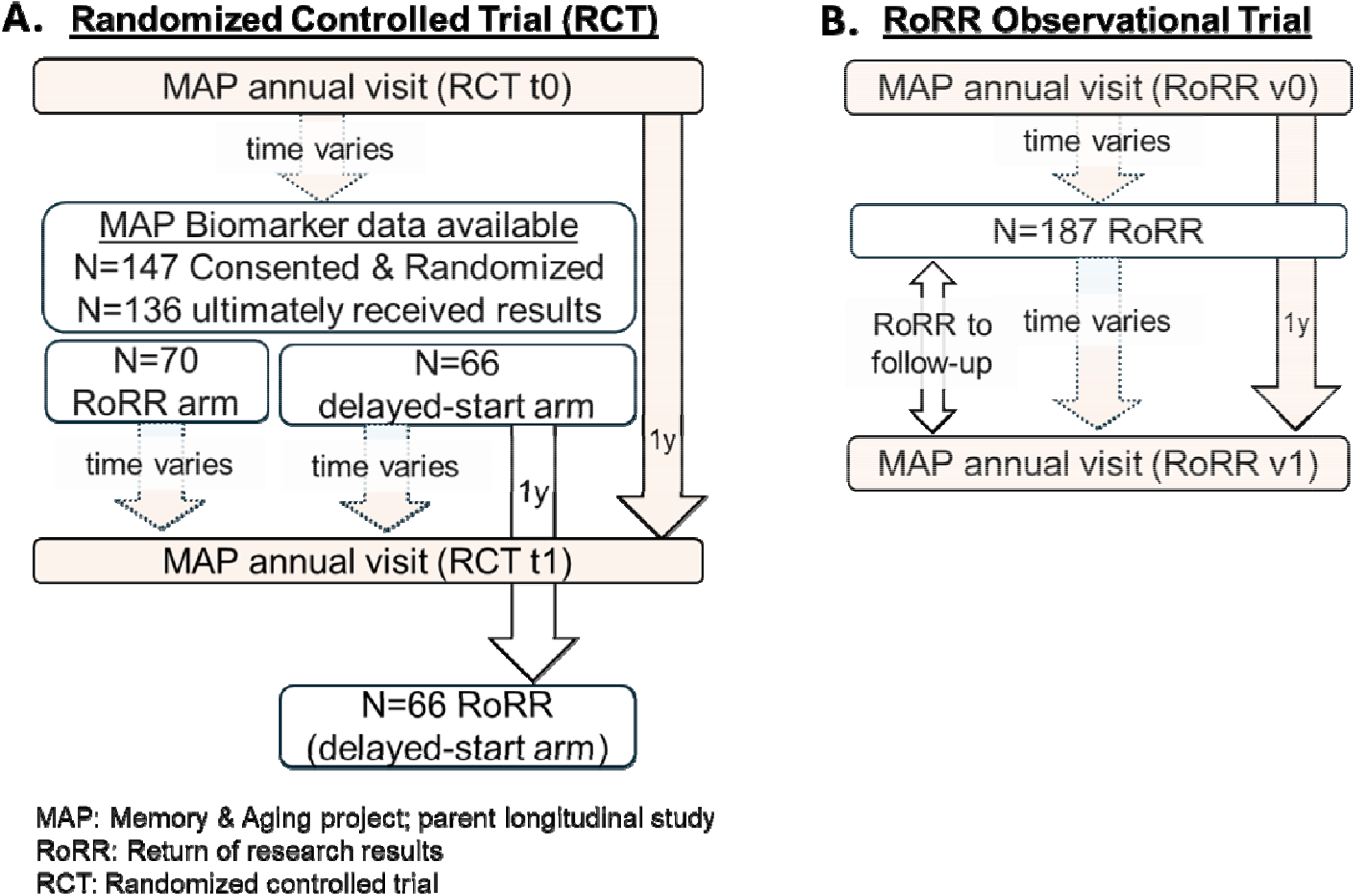
Study flow diagrams for randomized controlled trial (RCT A), and RoRR observational study (RoRR B). For RCT, primary outcomes are based on changes from RCT t0 to RCT t1. For the observational RoRR study, primary outcomes are based on changes from RoRR v0 to RoRR v1. All participants in the RCT are also part of the observational trial. For participants randomized to the RoRR arm, RoRR v0 and v1 correspond to RCT t0 and t1. For participants randomized to the delayed-start arm, RoRR v0 corresponds to RCT t1.

### Randomization

Participants were randomized 1:1 to RoRR either 2-4 weeks (intervention/RoRR group) or 1 year (control/delayed-start group) after consent. A randomization algorithm was used to preferentially balance the following factors: amyloid positivity, arm size, race, and gender. Because amyloid positivity was not known prior to randomization, family history was used as a proxy. The randomization parameters were set after running simulation studies. Details regarding the randomization procedures were previously described.^15^

A third group of participants received results but were not randomized because either study procedures needed to be piloted, or the participant was unwilling or unable to receive results 1 year later. Like the RoRR group, these participants received results 2-4 weeks after consent.

### Research Results Returned

Participants were offered their *APOE* genotype and either their imaging results (amyloid PET centilioid (CL) quantification and MRI hippocampal volume) or their plasma amyloid results (APS (amyloid positivity score) derived from plasma Aβ42/40, age and apolipoprotein E proteotype). Biomarker collection and processing has been described previously.^15,18–20^ Participants were given a report that included (1) a baseline absolute 5-year risk of AD dementia and their biomarker adjusted risk using our previously published algorithm,^18^ and (2) the individual results of the biomarker tests (APOE genotype, amyloid PET positivity, hippocampal volume low, plasma amyloid positivity based on APS).^15^ Results were returned either in-person or via Zoom HIPAA with a trained study clinician (MD, genetic counselor, or genetic counseling student) using a standardized approach. Participants were called 1 week later to answer any questions or concerns. Participants with questions that could not be satisfactorily answered by a staff member were offered either a telephone or Zoom follow-up appointment with a clinician.

### Outcomes

Primary outcomes were longitudinal change in symptoms of depression and cognition, measured at the annual MAP visits (**Figure 1**). Symptoms of depression were measured using the Geriatric Depression Scale (GDS).^21^ Cognition was measured in 2 ways: clinical measurement of cognition based on CDR sum of boxes score (CDR-SB),^16^ and objective measurement of cognition based on a global cognitive composite score.^22^ Clinicians involved in the outcome assessments were blind to the participant’s biomarker results. Outcomes were evaluated in the full sample and stratified by amyloid status. A secondary outcome was subjective change in memory, as defined by GDS item 10,^21^ “Do you feel that you have more problems with memory than most?” These measures were collected through the parent longitudinal MAP cohort. We added subjective memory impairment as an outcome of interest because more subjective impairment has been previously seen among participants who receive results indicating increased risk of AD.^13^ Additional WeSHARE-specific measures evaluated AD concerns, decisional regret, social impact, behavioral and lifestyle changes, and will be reported in another manuscript.

### Statistical analyses

Our primary analyses evaluate *longitudinal changes* in symptoms of depression (GDS) and cognitive function (CDR-SB and global cognitive composite) by comparing these measures from the MAP annual assessment preceding WeSHARE consent to the measures from the MAP assessment after WeSHARE consent (after RoRR for RoRR arm, before RoRR for delayed-start arm). This allows evaluation of the impact of RoRR on these longitudinal measures.

The primary analyses are non-inferiority tests of whether longitudinal change in symptoms of depression or cognition is worse for the RoRR arm relative to the delayed-start arm. This corresponds to a one-sided hypothesis test of increased change in GDS, increased change in CDR-SB, and decreased change in psychometric composite. The Shapiro-Wilk test was used to evaluate whether the outcomes were normally distributed. Difference in means of outcomes that were normally distributed were statistically compared using one-sided t-tests. Differences in outcomes distributions that were not deemed normally distributed based on the Shapiro-Wilk test were compared using a one-sided Mann-Whitney U test. Non-parametric 95% confidence intervals of the arm differences for the non-normally distributed outcomes were computed by bootstrapping with N=10,000 replications using the R package boot.^23,24^

Secondary analyses explored alternative coding of the primary outcomes, subgroup analyses, and analyses using secondary outcomes. Alternative coding of outcomes included chi-square analyses of whether the randomized arms differed with respect to the primary outcomes, coded dichotomously (“worse” vs “not worse”). Fisher’s Exact test was used for cell sizes <5. Subgroup analyses included evaluating individuals who were biomarker positive (elevated amyloid vs. not elevated amyloid), and excluding participants who had not previously had cognitive testing.

Finally, we tested the hypothesis that there is a time-dependent effect of RoRR on the outcomes by evaluating the association between RoRR and its proximity to the follow-up test for all participants. Because the consent visit was scheduled based on the availability of biomarkers, the proximity between consent and the follow-up measure was variable, averaging 6 months, but ranging from 0 – 14 months. This included both randomized and non-randomized participants and compared the assessment following RoRR to the assessment preceding RoRR.

All statistical analyses used SAS 9.4^25^ or R version 4.5.3.^26^ Statistical significance was defined as p<0.05. Clinically meaningful worsening for the primary outcomes were defined as a 4 unit increase in GDS, corresponding to a shift in clinical categorization from subclinical depression to mild depression; a 0.5 unit increase in CDR-SB, corresponding to a shift from normal function to slight impairment in a single cognitive domain (memory, orientation, community affairs or home & hobbies); and a standard deviation of average change in the global psychometric composite, as measured among participants who did not have elevated amyloid, corresponding to a Cohn’s *d* of 0.5.

Our primary analyses excluded missing data. Missing data may be either random or non-random. In secondary analyses we imputed missing values for the change in the primary outcomes by running a linear regression using baseline measurements of age, sex, educational attainment, parental history of AD dementia, and amyloid positivity to determine whether missingness may impact the study results.

## Results

Of 341 eligible participants from the MAP cohort, 200 consented. A total of 147 were randomized: 73 to the RoRR arm (immediate return, 70 received results) and 74 to the delayed-start arm (return 1 year later, 66 received results), **Figure 1A**. No adverse events were observed. Seventy RoRR and 66 delayed-start participants received results. The remaining 53 consented participants were not randomized due to pilot participation or specific disclosure needs; 51 received results (**supplementary Figure 1**). These participants were added to the RCT participants for an observational study of the impact of RoRR on longitudinal outcomes, **Figure 1B**.

Baseline demographics and clinical characteristics of randomized participants who received results were stratified by study arm and amyloid status (**Table 1**). The randomization algorithm ensured no differences in demographics or biomarkers between arms. However, one baseline discrepancy occurred among amyloid-positive (Aβ+) participants: those in the delayed-start arm had higher baseline symptoms of depression than those in the RoRR arm.

**Table 1:** Characteristics of the data: randomized participants who received results. Follow-up measures are post-RoRR for RoRR arm and pre-RoRR for delayed-start arm.

|  | Amyloid + |  | Amyloid - |  |
| --- | --- | --- | --- | --- |
|  | RoRR arm<br>(N=16) | Delayed-start arm<br>(N=21) | RoRR arm<br>(N=45) | Delayed-start arm<br>(N=54) |
| **Average age (SD)** | 80.6 (5.9) | 76.0 (4.9) | 72.2 (4.9) | 72.9 (4.5) |
| **Gender** |  |  |  |  |
| female | 9 | 10 | 25 | 33 |
| male | 7 | 11 | 20 | 21 |
| **Race** |  |  |  |  |
| Black | 0 | 0 | 6 | 3 |
| White | 16 | 21 | 38 | 51 |
| Other race | 0 | 0 | 1 | 0 |
| **Average years of education (SD)** | 16 (2) | 17 (3) | 17 (2) | 17 (2) |
| **Family history** |  |  |  |  |
| Known AD in parent(s) | 4 | 10 | 22 | 22 |
| No known AD in parents | 12 | 11 | 23 | 32 |
| **# APOE e4 alleles** |  |  |  |  |
| 0 | 11 | 9 | 35 | 40 |
| 1 or 2 | 5 | 12 | 10 | 14 |
| **Biomarkers offered** |  |  |  |  |
| Imaging | 7 | 15 | 40 | 47 |
| Plasma | 9 | 6 | 5 | 7 |
| **Months from consent to follow-up testing (SD)** | 6.6 (3.5) | 5.5 (4.5) | 5.7 (3.6) | 5.8 (3.0) |
| **Baseline measures** |  |  |  |  |
| **GDS score (95% CI)** | 0.60 (0.8) | 1.56 (1.5) | 1.13 (1.6) | 0.68 (1.3) |
| No symptoms (GDS 0) | 9 (56%) | 5 (24%) | 24 (44%) | 24 (53%) |
| Subclinical symptoms (GDS 1-5) | 6 (38%) | 12 (57%) | 20 (37%) | 16 (36%) |
| Depression evaluation indicated (GDS>5) | 0 (0%) | 1 (5%) | 1 (2%) | 1 (2%) |
| Missing | 1 (6%) | 3 (14%) | 9 (17%) | 4 (9%) |
| **CDR-SB score (95% CI)** | 0.03 (0.1) | 0.03 (0.1) | 0.00 (0.0) | 0.01 (0.1) |
| Missing | 0 | 2 | 3 | 8 |
| **Global cognitive composite (95% CI)** | 0.23 | 0.32 | 0.37 | 0.47 |
| Missing | 0 | 2 | 4 | 8 |
| **Subjective memory impairment** |  |  |  |  |
| no | 13 | 14 | 42 | 40 |
| yes | 2 | 4 | 3 | 1 |
| Missing | 1 | 3 | 9 | 4 |

Primary non-inferiority analyses evaluated longitudinal changes in depressive symptoms (GDS), clinician-measured cognition (CDR-SB), and objective cognitive performance (cognitive composite) by comparing measures from the MAP annual assessment preceding WeSHARE consent to the subsequent annual assessment. Longitudinal changes did not differ between arms overall. Because a participant’s response to research results likely differs for those who receive Aβ+ results rather than Aβ-results, we focused on outcomes stratified by amyloid status (**Table 2, Figure 2**). For all stratified analyses, confidence intervals fell within clinical significance bounds, supporting non-inferiority.

**Table 2:**
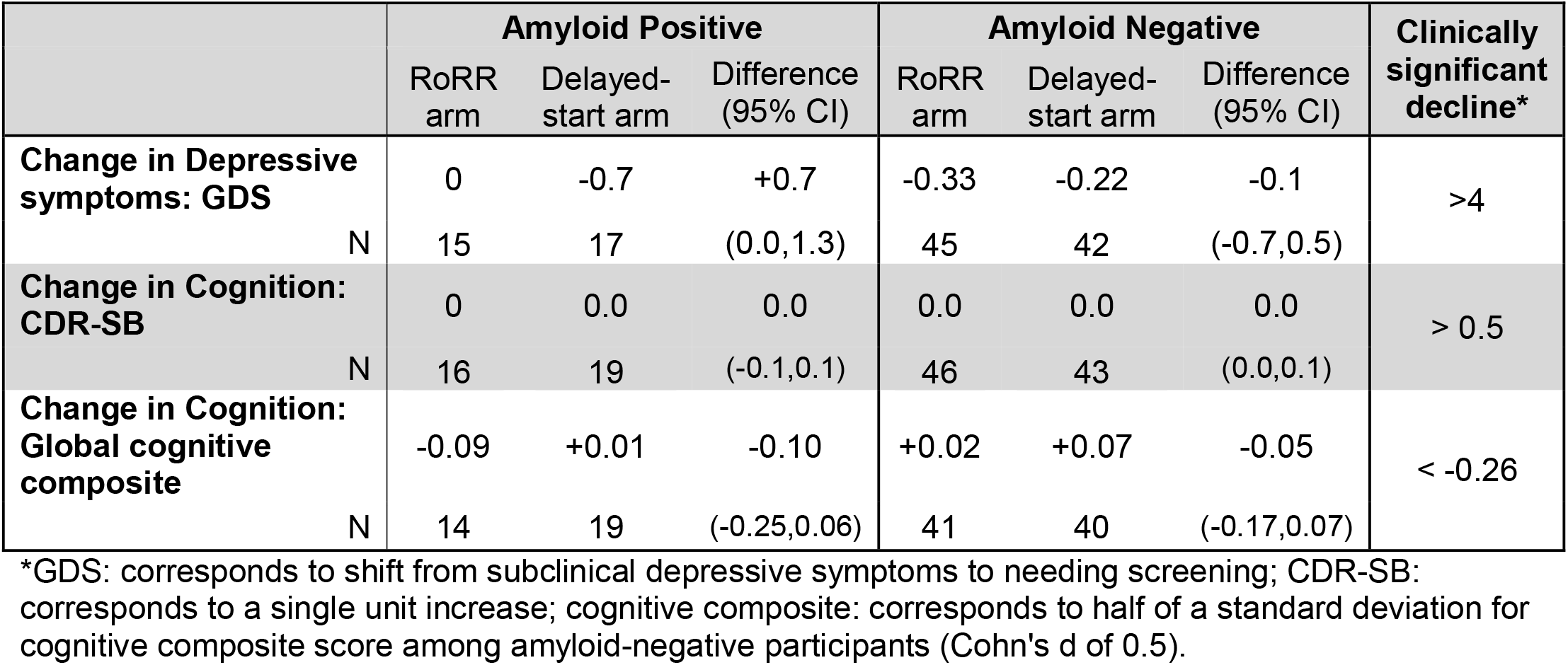
Changes in primary outcomes pre- and post-WeSHARE enrollment. No statistical differences between study arms were seen.

**Figure 2:**
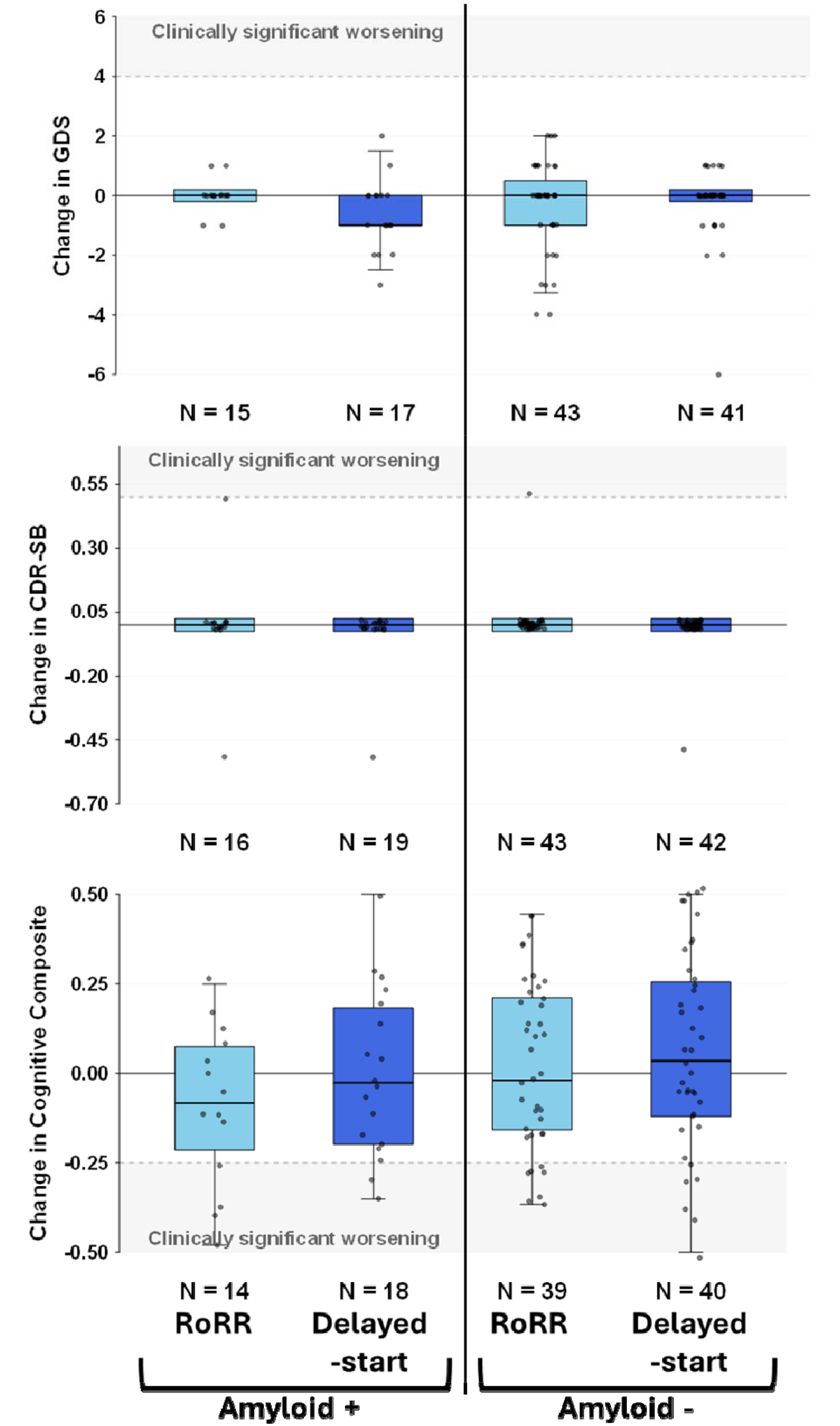
Box plots representing changes in primary outcomes for the delayed-start clinical trial, stratified by whether the participant had elevated amyloid. Light blue boxes represent changes for RoRR arm (before and after receiving results), and darker blue boxes represent changes in annual measurements prior to receiving results. The boundaries of the box denote upper and lower quartiles, horizontal line in the box is the median and error bars represent 95% confidence intervals. Clinically significant worsening is indicated by shading.

For GDS, longitudinal changes did not differ by arm in either the Aβ+ or Aβ-groups, with confidence intervals remaining below the clinically meaningful worsening threshold (GDS score change >4.0). Among Aβ+ participants, the average number of depressive symptoms decreased in the delayed-start group (average change of −0.7) and remained unchanged in the RoRR group (average change of 0), with a difference between the two arms of 0.7 (95% CI 0-1.3). This statistical difference (p=0.03) reflects the delayed-start group’s elevated baseline symptoms; however, the confidence interval falls well below the cutoff for clinical significance (GDS change of 4) suggesting that any statistical difference is not clinically meaningful.

For clinical cognition (CDR-SB), changes were identical across groups and confidence intervals fell within the non-inferiority bound (clinically significant decline defined as >0.5). The mean difference between arms was 0.0 for both the Aβ+ and Aβ-subgroups. The global cognitive composite score did not differ between groups and stayed within the threshold for clinically significant decline (< −0.26). The mean difference between arms was −0.10, 95% CI (−0.25,0.06), for the Aβ+ subgroup and −0.05, 95% CI (−0.17,0.07) for the Aβ-subgroup.

We ran several sensitivity analyses. Because changes in cognitive test results are impacted by whether the participant previously took the test,^27^ we ran the analysis excluding the 12 participants whose first exposure to the cognitive testing was the baseline test, and no substantive differences were seen. Finally, we used the demographic variables to impute the missing outcome values, and the model estimates did not substantively change.

Previous studies have found that response to RoRR wanes over time. Because our study design had variable length of time between RoRR and follow-up testing (ranging 0-14 months), we evaluated this using all 187 participants who received results and tested whether disclosure proximity to follow-up testing (time between RoRR and RoRR v1) was associated with longitudinal changes in outcomes. Linear regression modeling demonstrated that the number of months between disclosure and subsequent testing was not associated with longitudinal changes in depressive symptoms, CDR-SB, global cognitive composite, or subjective memory impairment. This lack of association was consistent across both Aβ+ and Aβ-subgroups (**Table 3, supplemental Figure 1**).

**Table 3:** Characteristics of the data: all participants who received results (including both randomized participants included in Table 1 and non-randomized participants).

|  |  | Amyloid elevated<br>(N=60) | Amyloid not<br>elevated<br>(N=127) |
| --- | --- | --- | --- |
| Average age (SD) |  | 78.4 (5.6) | 72.8 (5.2) |
| Gender | female | 29 (48%) | 74 (58%) |
|  | male | 31 (52%) | 53 (42%) |
| Race | Black | 0 (0%) | 12 (9%) |
|  | White | 60 (100%) | 114 (90%) |
|  | Other race | 0 (0%) | 1 (1%) |
| Years of education |  | 16.7 (2.5) | 17.0 (2.2) |
| Family history | AD in parent(s) | 26 (43%) | 62 (49%) |
|  | no known AD in parents | 34 (57%) | 65 (51%) |
| # APOE e4 alleles | 0 | 31 (52%) | 98 (77%) |
|  | 1 or 2 | 29 (48%) | 29 (23%) |
| Biomarkers offered | Imaging | 25 (42%) | 100 (79%) |
|  | Plasma | 35 (58%) | 27 (21%) |
| Average months from RoRR to follow-up (SD) |  | 6.2 (4.0) | 5.8 (3.7) |
| **Pre-RoRR measures** |  |  |  |
| Depressive symptoms: GDS |  |  |  |
| Average (SD) |  | 1.2 (1.4) | 0.9 (1.6) |
| No symptoms (0) |  | 36 (60.0%) | 87 (68.5%) |
| Subclinical symptoms (1-5) |  | 20 (33.3%) | 23 (18.1%) |
| Evaluate for clinical depression (>5) |  | 0 | 0 |
| Missing |  | 4 (7%) | 17 (13%) |
| Cognition: CDR-SB |  |  |  |
| Average (SD) |  | 0.1 (0.3) | 0.0 (0.1) |
| 0 |  | 54 (90.0%) | 125 (98.4%) |
| 0.5 |  | 6 (10.0%) | 2 (1.6%) |
| Missing |  | 0 | 0 |
| Cognition: Global cognitive composite |  |  |  |
| Average (SD) |  | 0.2 (0.5) | 0.4 (0.5) |
| Missing |  | 0 (0%) | 1 (0.8%) |
| Cognition: subjective memory impairment |  |  |  |
| No |  | 46 (77%) | 103 (81%) |
| Yes |  | 10 (17%) | 7 (6%) |
| Missing |  | 4 (7%) | 17 (13%) |
| **Change from pre-RoRR to post-RoRR** |  |  |  |
| Depressive symptoms: GDS |  |  |  |
| Average (SD) |  | -0.1 (1.1) | -0.1 (1.2) |
| worse (>0) |  | 6 (10.0%) | 20 (15.7%) |
| | not worse ($\leq 0$) | 32 (53.3%) | 63 (49.6%) |
|  | Missing | 22 (36.7%) | 44 (34.6%) |
| Association with proximity to RoRR* (95% CI) |  | -0.01 (-0.10,0.07) | 0.04 (-0.03,0.12) |
| Cognition: CDR-SB |  |  |  |
|  | Average (SD) | 0.10 (0.4) | 0.01 (0.1) |
| | worse ($>0$) | 2 (1.6%) | 6 (10.0%) |
| | not worse ($\leq 0$) | 125 (98.4%) | 54 (90.0%) |
|  | Missing | 0 | 0 |
| Association with proximity to RoRR* (95% CI) |  | 0.00 (-0.03,0.03) | 0.00 (-0.01,0.005) |
| Global cognitive composite |  |  |  |
|  | Average (SD) | -0.1 (0.2) | -0.0 (0.2) |
| | worse ($<0$) | 33 (55%) | 61 (48%) |
| | not worse ($\geq 0$) | 23 (38%) | 51 (40%) |
|  | Missing | 4 (7%) | 15 (12%) |
| Association with proximity to RoRR* (95% CI) |  | -0.01 (-0.02,0.01) | -0.01 (-0.02,0.003) |
| Cognition: subjective memory impairment |  |  |  |
|  | worse (unimpaired to impaired) | 1 (2%) | 2 (2%) |
|  | unchanged | 34 (57%) | 77 (61%) |
|  | improved (impaired to unimpaired) | 3 (5%) | 4 (3%) |
|  | Missing | 22 (37%) | 44 (35%) |
| Association with proximity to RoRR* (95% CI) |  | -0.01 (-0.03,0.01) | -0.01 (-0.02,0.004) |

\*Based on a linear regression of outcome as a function of months between RoRR and follow-up testing

## Discussion

We tested the impact of RoRR on longitudinal changes in depressive symptoms and cognition by randomizing participants to either receiving results 2-3 weeks after study enrollment or receiving results 1 year later (after follow-up testing). We found that returning AD biomarker results does not negatively impact longitudinal measures of depressive symptoms or cognition, both overall and in subgroups defined by whether the participants had biomarkers suggesting elevated amyloid. This study is the first to evaluate the impact of RoRR on longitudinal measures of cognition, the first to randomize return of amyloid results, and the first to return multiple biomarkers. The results support mounting evidence that returning AD research results to cognitively unimpaired older adults neither harms them nor significantly impacts longitudinal measures of cognition.

Using a randomized, non-inferior trial design we estimated the impact of RoRR on the change in depressive symptoms and cognition before and after study enrollment, allowing for a direct evaluation of the impact of RoRR on participants who chose to receive research results. Randomization after the decision to receive results allows for a direct comparison of the impact of RoRR on participants, which is important given a significant proportion of participants who say they want results do not ultimately decide to receive results.^17^ Based on 95% confidence intervals, the changes in all outcomes did not reach a clinically meaningful threshold, which is the benchmark for non-inferiority, both overall and in analyses stratified by amyloid positivity, suggesting that any impact of learning AD biomarker research results is not clinically meaningful. In particular, the confidence intervals highlight that the study is adequately powered to detect clinically meaningful outcomes, were they to exist. This confirms and extends the results from previous studies showing lack of clinically significant psychological harms, and provides first of its kind data on cognitive outcomes after AD biomarker disclosure.^6–8,10,11^ The lack of significant impact on longitudinal measures signifies that RoRR does not appear to impact key research study measures themselves, which is particularly important in light of heightened desire and expectation for RoRR in AD research studies.^1,2^

Prior studies have shown that effects of RoRR may decrease over time.^4,6,10,14,28^ To test this hypothesis, we evaluated whether there was an association between proximity to follow-up testing and longitudinal changes in depressive symptoms or cognition. No association was observed in either participants receiving Aβ+ results or participants receiving Aβ+ results. This suggests that RoRR can occur independently of study assessments, without concern that RoRR may impact symptoms of depression or cognition if it is too close to the study assessment.

The major limitation of this study is that our participants are mostly White and highly educated, limiting the generalizability of the study and leaving evidence gaps about the impact in more diverse individuals.^6,9,29–31^ Given that under-represented groups, such as Black and Latino individuals, are at elevated risk for AD,^32^ expansion to include these groups in disclosure studies is crucial. In addition, because we explicitly evaluated the impact on longitudinal changes in symptoms of depression and cognition, the results may not generalize to other psychological or behavioral factors. Fortunately, other outcomes are evaluated in a companion paper, which will help improve our understanding of the nuances of the impact of RoRR.^33^

Despite these limitations, our findings provide compelling evidence that, overall, there are no negative psychological or cognitive harms due to RoRR in cognitively unimpaired individuals. Further studies will need to evaluate the frequency of possible rare negative events within amyloid positive subgroups. While monitoring should continue to evaluate the safety and the effectiveness and clarity of the RoRR process particularly among underrepresented participants, the time has come to move beyond concerns for psychological harm and shift towards evaluating disclosure methods and ensuring scalability. With no evidence of harm, we urge researchers to take the leap towards disclosure, which is becoming an ethical obligation rather than source of ethical concern.

## Data Availability

Data collection is available via the Knight ADRC. Knight ADRC data can be requested through the online request portal on the Knight ADRC website: https://knightadrc.wustl.edu/professionals-clinicians/request-center-resources/.

## Acknowledgment

We would like to thank the Knight ADRC participants for their ongoing contributions to the project, and to Levi Levin for assistance with data collection in the first 2 years of the study. Sarah Hartz and Jessica Mozersky, Multiple Principal Investigators of the study, had full access to all the data in the study and take responsibility for the integrity of the data and the accuracy of the data analysis.

## Funding

R01 AG065234 (Hartz and Mozersky), R01AG070941 (Schindler), R01AG067505 (Xiong), P30 AG066444 (Holtzman DM) P01 AG003991 (Morris, JC), P01 AG026276 (Morris, JC), from the National Institute on Aging; UL1 TR002345 (Powderly, W) from the National Center for Advancing Translational Sciences.

## Role of Funder

The funder did not have any role in the design and conduct of the study; collection, management, analysis, and interpretation of the data; preparation, review, or approval of the manuscript; and decision to submit the manuscript for publication.

## Data Sharing

Data is available via the Knight ADRC. Knight ADRC data can be requested through the online request portal on the Knight ADRC website: https://knightadrc.wustl.edu/professionals-clinicians/request-center-resources/.

## Disclosures

SMH received speaking fees from Novo Nordisk. JH is a paid consultant for Abbvie, Quanterix, and Prothena. SES has not received any financial compensation from pharmaceutical or diagnostics companies since 2024, when she received honoraria for serving on scientific advisory boards on biomarker testing and education for Eisai and Novo Nordisk and speaking fees from Eisai, Eli Lilly, and Novo Nordisk. She has recently received honoraria for educational presentations from Medscape, PeerView, and the Academy for Continued Healthcare Learning. She has provided unpaid scientific advising to Acumen, Biogen, Cognito Therapeutics, Danaher, Eisai, Eli Lilly, Johnson and Johnson Innovative Medicine, Sanofi, and Siemens. TB participates as a site investigator in clinical trials sponsored by Eli Lilly and Company, Biogen, Eisai, Janssen, and Roche. TB has been a consultant (paid and unpaid) to Eli Lilly and Company, Biogen, Eisai, Siemens, and Janssen. TB has received in-kind research support from Hyperfine, Lilly, Lantheus, and Life Molecular Imaging.

The study principal investigators (JM and SMH) would like to report that the parent clinical trial from which this data was drawn includes returning results from C2N diagnostic’s PrecivityADTM plasma AD biomarker test to participants who opt to receive their results. Washington University has a financial investment in C2N diagnostics, such as stock or other equity interest. Study activities have been reviewed by Washington University’s Institutional Conflict of Interest Committee (ICOIC) in accordance with WU’s Institutional Conflict of Interest (ICOI) Policy. No individual involved in the conduct of this study has a personal financial interest in C2N diagnostics. This information is contained within the informed consent documents for the study.

The other authors declare no Competing Financial or Non-Financial Interests.

---

**Supplementary Figure 1:**
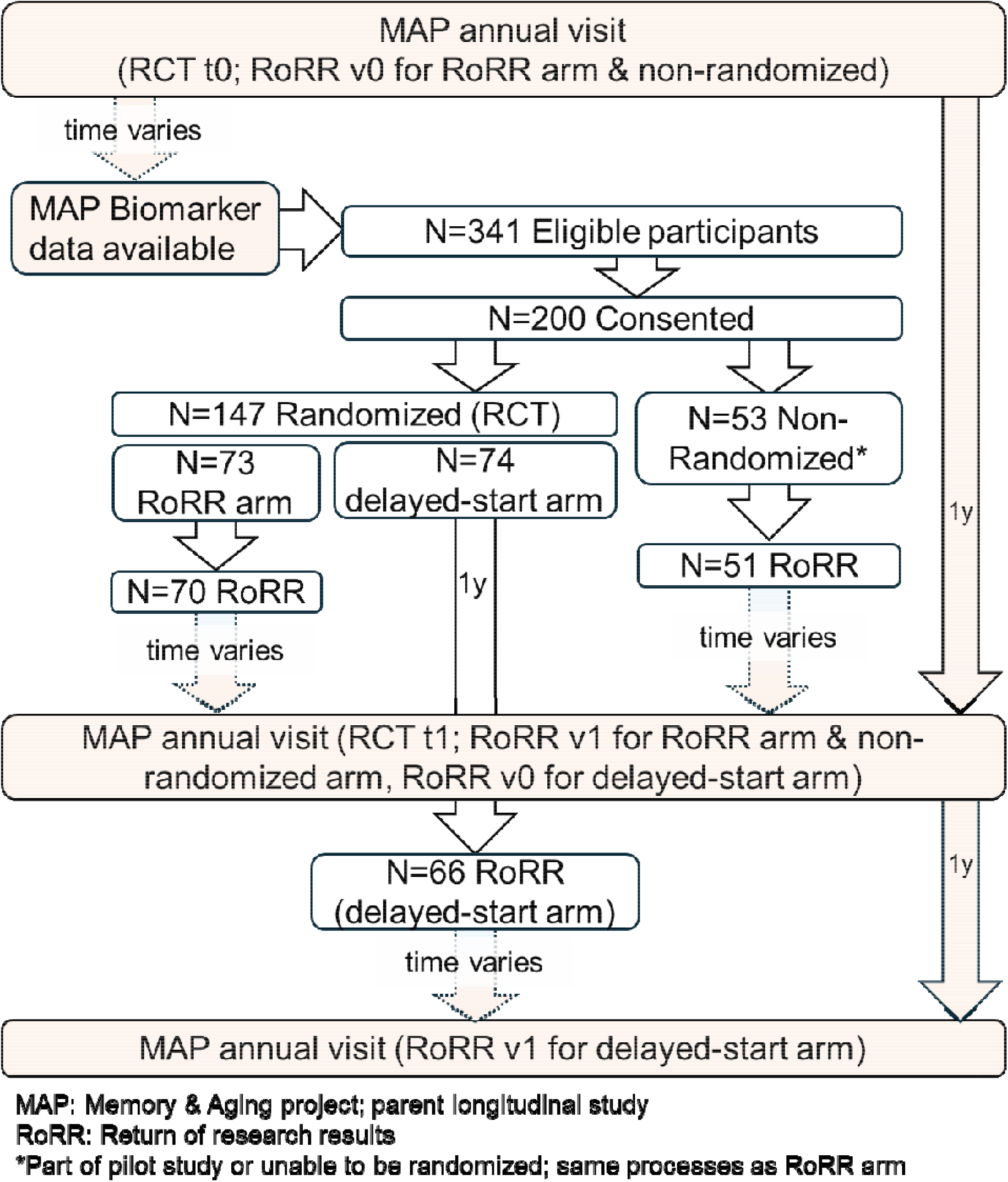
Integrated WeSHARE study flow diagram. MAP:Memory & Aging project; parent longtudlnal study RoRR:Return of research results *part of pilot study or unable to be randomized; same processes as RoRR arm

**Supplementary Figure 2:**
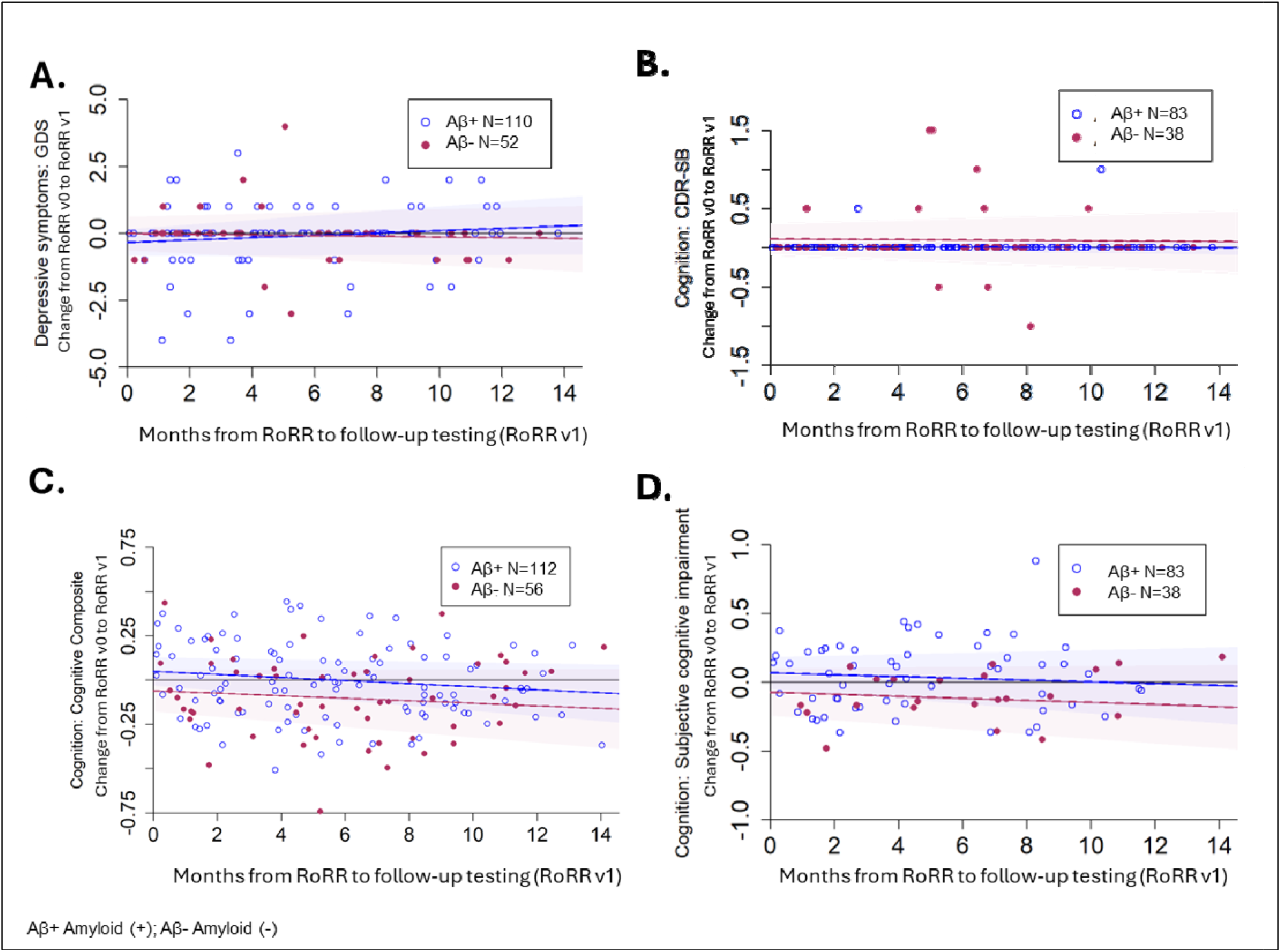
Observed changes in outcome based on proximity from RoRR to follow-up outcome measurement GDS (A), CDR-SB (B), cognitive composite (C), subjective cognitive impairment (D).

## Notes

### Clinical Trial

NCT04699786

### Author Declarations

The Institutional Review Board at Washington University in St. Louis gave ethical approval for this work

### Summary of Updates

This version has been edited to comply with journal requirements

